# Virtual care and the impact of COVID-19 on nursing: A single centre evaluation

**DOI:** 10.1101/2021.06.03.21258276

**Authors:** Luke Hughes, Anika Petrella, Natasha Phillips, Rachel M Taylor

## Abstract

**Aims:** The overall aim of this evaluation was to look at the impact of the changes in working practices during the pandemic on nurses. This secondary analysis provided an evaluation of virtual care and being able/required to work from home.

**Design:** This was secondary analysis of an evaluation using semi-structured interviews.

**Methods:** Conducted at a single National Health Service (NHS) university hospital in the United Kingdom between May-July 2020. Forty-eight operational leads and nurses participated in semi-structured interviews which were digitally recorded, transcribed verbatim and analysed using a framework analysis.

**Results:** Two overarching themes emerged relating to the patient experience and nursing experience. There were both positive and negative elements associated with virtual care and remote working related to these themes. However, the majority of nurses found virtual clinics were useful when proper resources were provided, and managerial strategies were put in place to support them. Participants felt virtual care could benefit many but not all patient groups moving forward, and that flexibility around working from home would be desirable in the future.

**Conclusion:** Virtual care and remote working were implemented to accommodate the restrictions imposed because of the pandemic. The benefits of these changes to nurses and patients support these being business as usual. However, clear policies are needed to ensure nurses feel supported when working remotely and there are robust assessments in place to ensure virtual care is provided to patients who have access to the necessary technology.

**Impact:** This was a study of the move to virtual care and remote working during the COVID-19 pandemic. Telemedicine and flexible working were not common in the NHS prior to the pandemic but the current evaluation supports the role out of these as standard care with policies in place to ensure nurses and patients are appropriately supported.

## INTRODUCTION

In the era of the novel coronavirus (COVID-19) there have been huge changes to everyday life, effectively altering how we currently approach healthcare (Wosik et al, 2020). In the spring of 2020 the Government in the United Kingdom (UK) implemented national and regional lockdowns to minimise the rate of community transmission and protect the National Health Service (NHS) as it attempted to cope with the virus. Due to the pathogenicity and virulence of COVID-19, face-to-face clinical appointments were greatly reduced, and outside of urgent trauma care, significant restrictions were placed on outpatient care to limit hospital footfall, reduce patient to clinician transmission and prevent the spread in the general community (Wosik et al, 2020). In order to respond to the new demands, virtual care quickly became a necessary surrogate to in-person care (Bashshur & Shannon, 2020, Murphy et al, 2020). Virtual care reflects a spectrum of interactions between patients and/or members of their healthcare team delivered remotely, wherein the application of information and communication technologies are used to provide elements of healthcare without the need for face-to-face contact (Shaw et al., 2018, Speyer et al, 2018, Siegel, 2017).

## BACKGROUND

Virtual care has been in use throughout the last century, yet full scale adoption into healthcare systems has yet to be achieved (Bashshur & Shannon, 2009). Historically the medical community has been reluctant to fully engage with virtual care, and opinions on its efficacy have been mixed despite the evidence supporting its practicalities and use by a broad range of health professionals (Wosik et al, 2020). Prior to the COVID-19 outbreak, interest in virtual care was on the rise. In 2018 the World Health Organization (WHO) called on governments to assess the current/potential use of digital technologies in their healthcare systems (WHO, 2018). The NHS responded with a comprehensive digital transformation strategy (NHS England, 2019). However, it was the rapid onset of COVID-19-specific restrictions that became the main driver for immediate adoption of virtual care in the UK. Virtual care has the potential to address the on-going challenge of timely access to health care. For healthcare professionals, virtual care has been shown to provide greater flexibility in their working day, as well as improved autonomy in their provision of patient care (Hollander & Carr, 2020, Hoffmann et al, 2020). For patients, the use of virtual clinics reduces travel costs and has lowered overall admissions to hospitals in certain patient groups, such as the elderly (Lilliecrap, Hunter & Goldswain, 2019). The use of virtual care for some, if not the majority of healthcare appointments, can help provide equitable healthcare to more remote communities (Stokel-Walker, 2020, Wosick et al, 2020) and contributes to shorter wait lists, which are critical for patients with quickly deteriorating conditions or seeking a timely diagnosis (Murphy et al, 2020). Reducing waiting times consequently allows for higher volumes of patients to be seen by the appropriate professional, thus benefiting the system as a whole (Lilliecrap, Hunter & Goldswain, 2019).

The field of virtual care faces a number of critical challenges that require attention. Concerns have been raised specific to continuity of care, education and training of healthcare providers, and the potential risk of limited digital health literacy further exacerbating health inequalities (Narasimha et al, 2017). It is argued that virtual care is not suitable for all patients, for example those with complex needs, those who do not have access to or feel comfortable using technology (Narasimha et al, 2017, Wosik et al, 2020). Technological issues, such as poor quality or lagging of video feed can negatively impact the clinician’s ability to gauge body language and nonverbal cues and affect their ability to provide adequate consultations (Sinha et al, 2020). Thus, it is critical that virtual care be embedded as a complementary pathway to providing care where appropriate rather than fully replacing face-to-face delivery of health services.

Prior to the COVID-19 pandemic, patients and clinicians were hesitant to engage with virtual care and change well established routines (Lilliecrap, Hunter & Goldswain, 2019; Sharma & Clarke, 2014). Yet for those who did engage, satisfaction was high (Azad et al., 2012). During the COVID-19 pandemic, virtual care offered a way to balance the supply of clinical services during each surge in demand, while also providing healthcare access regardless of physical or geographical boundaries (RGCP, 2020). This further helped protect the available stock of important resources such personal protective equipment and enabled shielding patients to maintain communication with their healthcare team (Hollander & Carr, 2020). Early reports suggested high levels of satisfaction among those who engaged in virtual care in the UK during the pandemic, with 98% reporting a desire to use virtual care again, even after COVID-19 restrictions were lifted (RGCP, 2020, Sinha et al., 2020).

A secondary element of virtual care highlighted during the pandemic was the ability for healthcare professionals to work remotely. Remote working helps protect staff classed as high risk, such as those who were immunocompromised or caring for vulnerable dependants (Wosik et al, 2020). Working from home presents a number of challenges and has been met with a level of reservations from a mostly conservative workforce (Giurge & Bohns, 2020, Tawfik, Profit & Magenthaler, 2018). Recent studies indicate working from home is not only possible but effective, and in many cases, a preference for healthcare professionals if they are given the opportunity (Chattopadhyay, Davies & Adhiyaman, 2020, Hoffmann et al, 2020). However, negative experiences associated with working from home have also been reported, specifically around lack of separation between work and home life (Giurge & Bohns. 2020), difficulties caring for dependents (e.g. schools being shut resulted in balancing work with childcare), or issues with technology (Hoffmann et al, 2020). A study of all professional groups working in the NHS, who were working from home during the pandemic showed that 43.4% felt their work was undervalued or not acknowledged in comparison to their frontline colleagues, and 48% struggled with feelings of guilt due to being at home during a crisis (Chattopadhyay, Davies & Adhiyaman, 2020).

The integration of virtual care into standard practice is well on its way and embodies a new normal for healthcare after the resolution of COVID-19. In particular it is believed to be key to improving communication between healthcare, the patient and their wider systems, which has important implications for their treatment outcomes (Hollander & Carr, 2020). If the benefits of virtual care are to be fully realised within the NHS, a thorough understanding of individuals’ experiences using virtual care during this unique time is needed.

## THE STUDY

### Aim

The purpose of this study was to explore nurses’ experiences of utilising virtual care, alongside remote working to identify what elements could be implemented into a recovery model following the conclusion of the pandemic.

### Design

This was secondary analysis of semi-structured interview data collected from nurses as part of a wider service evaluation of the changes to delivery of care to accommodate the pandemic, at a single university hospital in the UK.

### Participants

An initial purposive sample of hospital-wide operational leads were recruited through targeted invitations from a senior nurse, to describe the changes to services across the hospital (n=17), then a convenience sample of nurses at different levels of seniority were invited to participate through the Trusts group email lists (n=31). This secondary analysis focused on the experiences of matrons (n=7), sisters/charge nurses (n=8), clinical nurse specialists (CNS; n=14) and clinical research nurses (CRN; n=2).

### Ethical considerations

The evaluation was conducted in accordance with the UK Framework for Health and Social Care Research (Health Research Authority, 2017). The purpose of the evaluation was explained to participants at the beginning of the video call, who then were given the opportunity to ask questions. If they were happy to continue, they were asked to give a recorded consent. All participants were able to stop the interview at any time and were assured of anonymity and confidentiality.

### Data collection

Data were collected through individual semi-structured interviews between May and July 2020. The guide for the interviews with operational leads included a description of the changes in service delivery the participant led and their perception of what worked well and what were the challenges. The analysis of these data informed the structure of the interviews with nurses, reflecting on their experiences of the service changes and what they felt worked well or could be improved. Interviews were conducted through video conference software and were digitally transcribed.

### Data analysis

Digitally recorded interviews were transcribed verbatim and analysed using Framework Analysis (Richie and Spence 1994). The current evaluation analysis focused on the aspects of the Framework that referred specifically to virtual care and working from home. A secondary Framework was developed specifically in these areas to further illuminate the experience. Transcripts were re-reviewed and additional indexing applied from the new framework. The main framework was developed by two members of the evaluation team, checked by an independent researcher with expertise in qualitative research; the secondary framework by reviewed by a third member of the evaluation team.

### Rigour

The criteria proposed by Beck (1993) were used to establish methodological rigour. Credibility was established by using a semi-structured guide for the interviews but also empowering participants to expand on their responses according to their personal experiences. To ensure fittingness of the findings, the secondary analysis included a purposive sample of nurses whose practice was impacted by the pandemic to require remote working and the move to virtual clinics. To ensure the auditability of the findings, Framework Analysis was used, which enabled multiple researchers to review the coding to check for accuracy of the interpretation.

## FINDINGS

There were two overarching themes emerging from interviews: the perceived barring virtual care had on patient experience from a nursing perspective; and nurses’ experiences of virtual care and remote working. It was found that for most themes there existed a duality of positives and negatives.

### Potential barring on patient experience

This theme is based on nurses’ perceptions of how virtual care impacted their patients, which comprised of multiple subthemes (Figure 1).

**Figure 1.**
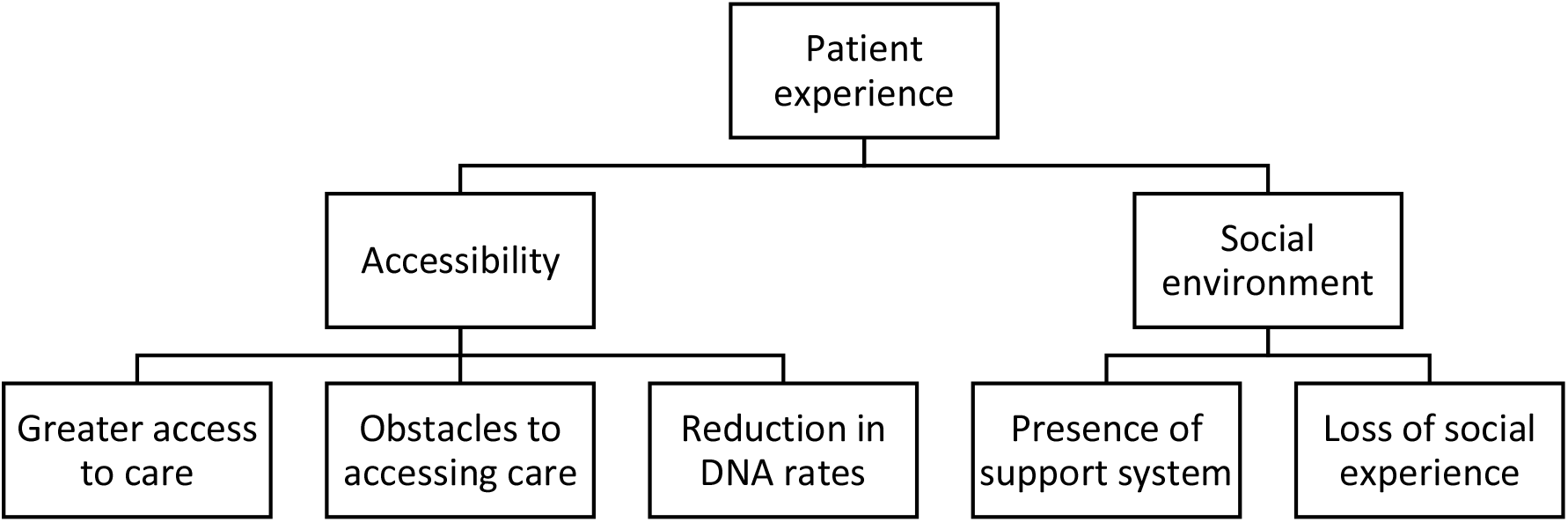
Major themes and subthemes for the potential barring on patient experience DNA: did not attend

#### Accessibility

##### Greater access to care

Participants identified areas in which they perceived the use of virtual care had positive impacts on patient’s experiences. In particular, the majority believed virtual care facilitated greater access to medical care for their patients, while also eliminating various barriers such as travel, finance and having to balance appointments with work schedules. By offering virtual appointments to patients, it allowed for them to attend remotely which was perceived as a benefit for many who would have to make long commutes into the hospital and eradicated their need to take time off of work to attend these appointments.

> *“Some of our patients live quite some distance away with, you know, they might have mobility issues or lots of other comorbidities that make getting to hospital difficult and expensive, of course. So I think I like the idea that we can offer them more choice at the moment,”* (CNS)

This also eliminated the process of having to return home after making a journey to the hospital, which could be difficult if the appointment involved bad news. This was particularly salient during the pandemic as heavy restrictions on visitor policies meant many would have had to attend clinics alone. Allowing patients to remain at home with family and access their clinics virtually therefore negated this potentially distressing circumstance.

> *“I think also a lot of patients have preferred not to come in, not doing face to face clinics, a lot of patients, you know, have found telephone clinics very helpful. You know, they’ve been protected, they’re home,”* (CNS)

##### Obstacles to accessing care

Despite the benefits of virtual care, nurses also perceived some negative impacts for their patients. One of the main difficulties was the lack of accessibility of technology for some patients, such as the elderly. Furthermore, for those who had access to technology, they did not necessarily have the capability to use it with confidence, and if they did not have support systems at home to help, this was a potential obstacle for patients to use it. Some nurses noted that there were certain subgroups of patients who were unable to engage at all with virtual appointments during the pandemic and feared that they were at risk of becoming neglected by these advances in technology. Similarly, technological issues inherent to these platforms could frustrate or further aggravate a patient’s reluctance to engage. Simple things such as the quality of the video could affect the flow and efficacy of the intervention.

> *“I do know that a lot of people, you know, a lot of patients may not have that capability. The other thing about that is…if there’s any technical problems, it can delay things hugely,”* (CNS)

##### Reduction in DNA rates

Nursing staff felt virtual care had clear benefits for protecting vulnerable patients from making unnecessary journeys during the pandemic, and as a result of this, the ‘did not attend’ (DNA) rates were noted to be lower than normal, as patients were able to attend easily without making too much compromise in their day-to-day lives. The convenience of not having to attend the hospital also had a financial benefit, as many patients were receiving specialist care so the hospital was not local. The use of virtual care appeared to create a positive feedback cycle in which remote access to their healthcare workers helped to facilitate their care, without much personal or financial cost to themselves.

> *“DNA rates…have been very, very low, you know, typically runs around 14% or so. I think probably it’s 5% if that, you know, because people are home usually, and seem to appreciate the call,”* (CNS)

Staff noted that due to the decrease in DNA rates they were able to see a much higher volume of patients than they would see in face-to-face clinics. Nurses felt this made them more productive and removed the necessity for rescheduling patients and delaying elements of their treatment. They also noted that the ability to remotely access patients negated the Trust’s need to organise costly travel for those who could not travel by conventional means, which was a further financial and time benefit for the NHS.

> *“Hospital transport, that must be costing the NHS a fortune, I’d be more inclined to say to them now, lets not go with the hospital transport and everything. Let’s do it by telephone,”* (CRN)

#### Social environment

##### Presence of support system

Another benefit highlighted by nurses was that it gave patients the opportunity to have more of their support systems present during their appointments. Prior to use of virtual clinics it could be logistically difficult for patients and their families to attend clinics together. This resulted in some patients attending clinics alone, lacking their support structures and having their family feel excluded in terms of medical developments. Likewise, families may struggle to attend appointments while balancing other life responsibilities. With the option of virtual clinics, patients and their family members could be more easily present, meaning the patient had the added benefit of being in comfortable surroundings with their support structures. Family members could be present to engage with nurses who could address any questions or concerns they had. In doing so this mutually strengthened the relationships between the patient, their support systems and the medical team.

> *“Their partner or family member can join in on the conversation. And certainly from speaking to my colleagues, we’ve all felt that and doing telephone clinics is a way forward,”* (CNS)

However, others felt that this may not suit everyone, and that there were patients whom they felt benefited from attending face-to-face clinics. This included patients where there were safeguarding concerns, or those who did not wish to have family involved in their care, and therefore finding space and privacy for virtual appointments was more difficult. Likewise, for those with children or dependants, patients felt uncomfortable discussing health issues around them, which became more difficult when appointments were mostly arranged at home.

> *“If I’m sitting at home, and I’ve got young children at home, I can’t express myself clearly, you know, openly to discuss everything,”* (CNS)

##### Loss of social experience

The inverse of the removal of travelling for appointments, seen by many as a positive outcome, for some patients this was one of their main social outlets. Vulnerable patients or those with complex needs may not get to engage socially as easily as others, particularly during times of social distancing or shielding. Some staff worried that this may isolate those patients further and could have potentially negative effects on their wellbeing. Likewise, for some patients where hospital visits were regular parts of their routine, the relationships they built with staff could be minimised by moving towards more virtual based care. Interestingly, this ran parallel to experiences of staff who were working at home remotely, realising how isolating it can be, and missing the everyday social connections with their co-workers.

> *“…sometimes it’s the only way that they get to leave where they live. That’s depriving them of that,”* (CNS)

### Nurse’s experiences of virtual care and remote working

This theme relates to using virtual care and remote working in the nursing role, and the ways in which it both positively and negatively affected nurses’ experiences. The subthemes are summarised in Figure 2.

**Figure 2.**
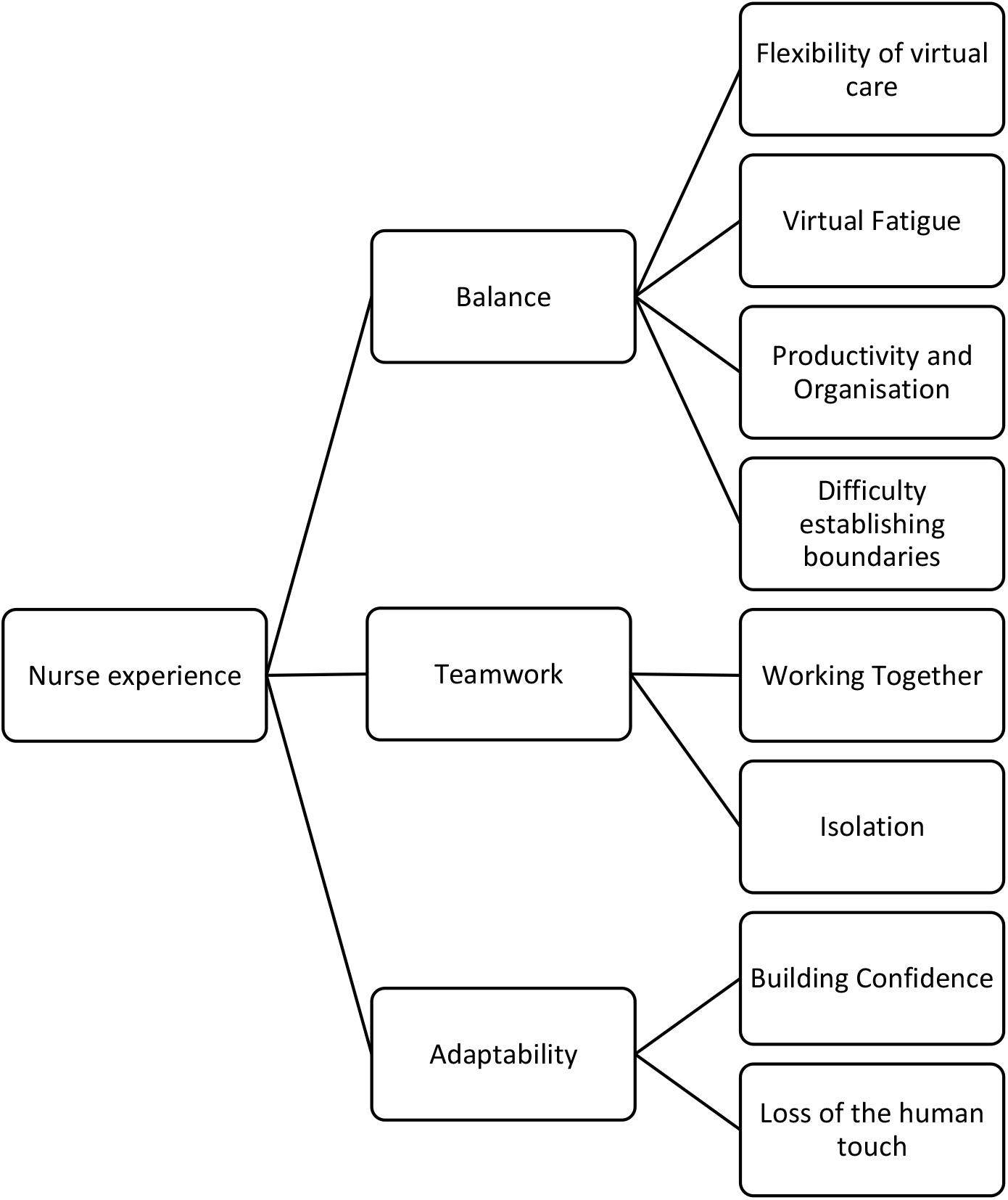
Major themes and subthemes for nurse’s experiences of virtual care and remote working

#### Balance

##### Flexibility of virtual care

The majority of nurses felt that virtual care was beneficial for their patients, and in turn beneficial for themselves. A key benefit of using virtual care for outpatient clinics was that they were more likely to run to time, which historically was difficult to achieve in everyday practise. Logistical issues, such as patients being late to clinic due to transport problems were negated. Long waiting times were greatly reduced, and with patients being able to wait for clinics in the comfort of their own homes this was perceived to be much less arduous then spending long amounts of time in hospital waiting rooms. Removing these unavoidable frustrations from clinic days allowed for greater flexibility for both staff and their patients and helped improve rapport and experience for both groups.

> *“One of one of the issues that we’ve always had is that our clinics never run to time. And what happens then, is that you get patients who eventually come in for their clinic, they’re very distressed, they can be very angry. So before you can even start talking about what they’re there to discuss, you have to kind of break down some of those barriers by trying to calm them down, you know, lots of apologies, that sort of thing. So the fact that these people aren’t actually making their way to the hospital spending, you know, extended waits in the [location] waiting for their appointment, the fact that they can do that at home and just be getting on with their daily lives. I think for everybody’s stress levels, doctors and patients and nurses and clinic staff, who were all patient facing, I think it makes it a lot less stressful for everybody,”* (CNS)

##### Virtual fatigue

Fatigue from attending large numbers of online clinics or meetings was noted, which participants felt required a different level of cognition to pay attention to, compared with face-to-face interactions, as they lacked much of the stimulation of in-person presence. Some felt there was a certain level of coldness associated with attending meetings online, particularly if those present did not turn on their cameras. This further reflects staff feeling isolated or cut off from their co-workers.

> *“I think meetings virtual meetings are much more productive, although I have to say they are exhausting. And I did six the other day. And I was absolutely exhausted because it’s all about concentration, isn’t it and listening, whereas if you’re in a meeting with a roomful of people, you can probably drift off a little bit or look at your emails or you know, you don’t have to so much concentrate on the conversation. Whereas if you’re doing a virtual meeting, you have to listen. Always, all the way through the meeting in order to capture what’s being said done, decided, etc,”* (CNS)

##### Productivity and organisation

Having the option to work remotely was noted by many nurses as having a positive impact on their work. They perceived it to be associated with higher levels of productivity and organisation. Nurses felt that working in the home environment allowed them to get much more of their paperwork completed than they would in the hospital setting, where they were often distracted.

> *“It has been proven that it was effective. And we managed to cover every single aspects of our job without compromising any single one. And it’s nice; we have a great routine, great organisation, etc, etc. So that’s the positive side of COVID that I can see…I find myself much more organized,” (CNS)*

Having the ability to set aside time to get these tasks done without distractions also equated with reduced feelings of stress. Remote working provided a comfortable and quiet environment free from the stressors of onsite work, allowing nurses to engage with tasks they often found difficult to apportion appropriate time to for.

> *“And I think because my mind is much clearer when I’m at home, there’s no distraction. So I get things done very quickly… and very effectively,”* (CRN)

Furthermore, organisational annoyances such as hot desking were essentially eradicated. In normal circumstances, desk space in the hospital was often an area of contention for nurses who were expected to do certain amounts of desk work. This was heightened during times of social distancing. Allowing nurses the option to work from home, for even some of the week, gave them the ability to process these tasks efficiently and therefore allowing them to engage more with clinical work when onsite.

> *“I can plan my day in a much better way …like we were fighting for computers in the office. There was no space for all of us to be, how many times I arrived in the office and I had to go back to the command centre because there was no desk…we were all hot desking”* (CNS)

##### Difficulties establishing boundaries

In terms or remote working, while many did find it greatly increased their productivity some felt it actually increased the level of pressure in their role. Some displayed levels of guilt at not being onsite during a crisis and made personal compromises such as a working extra hours or working the time they would have spent commuting– feeling they owed this to their colleagues. Others noted that when working from home they felt increased pressure from onsite colleagues to be able to complete tasks for them quickly, and that there was a perception and expectation that those working from home had more time to get these tasks done rapidly

> *“I’ve never spent so much time on the laptop, at home, and even at times, I would go overtime. After five…I can still find myself working, you know, and because they gave me a lot of worksheets to do, like different studies and have to create all of those, and I was really busy,”* (CRN)

The boundary between work and home life was sometimes difficult to establish. Some nurses noted that when they were working from home indefinitely it could lead to feelings of lower motivation. Lacking the normal everyday experiences of commuting to work, encountering colleagues and meeting patient face-to-face meant staff were less stimulated throughout the day. Some felt this lent itself to feelings of loneliness, lethargy and boredom, and that working from home meant the aspects of their life in which they used to relax or enjoy became synonymous with their working days.

> *“What’s been at times difficult psychologically is just not getting out the house and not having that kind of clear, clear-ish division between what’s work and what’s home. So, you know, I, I come up with stairs, I’m in a spare room/study, and that’s my commute to work. And when I’m finished work, straight out of, you know, doing patient stuff…I’m down in the kitchen with the rest of the family. And so, I guess in some ways…strangely, I miss the commute, because it gave me that period of that time and space between work and home,”* (CNS)

#### Teamwork

##### Working together

Another positive aspect of using remote access for staff included the use of virtual platforms for multidisciplinary team meetings (MDTs). Many nurses noted the benefits of being able to attend meetings virtually, which allowed for greater flexibility in their working day. MDTs were more likely to be attended by a wider selection of staff involved in patient care, which facilitated the perspectives of different disciplines to be voiced and interactions with one another on a more frequent basis. This helped nurses form a holistic perspective of patient care and was believed to benefit their approaches to treatment. Furthermore, it encouraged more interaction between disciplines which may not have had a chance to meet in person previously often due to conflicting schedules.

> *“I think having more different disciplines working together closely, looking after patients works really, really well. So I think that would be great to take that forward and having staff who don’t normally work on the front line or are more in the back sort of clinics or, or in labs, having them come out, and sharing their knowledge with us was really useful. And having them review patients with us as well, bringing their expert knowledge in was really helpful,”* (CNS)

##### Isolation

Despite the increase of interdisciplinary communication, nurses who were mostly working remotely voiced feeling isolated from their own teams and missed having the contact they would normally have with their colleagues in person. There was a sense of isolation and loneliness, which some described as feeling not only disconnected from their role and the work with their patients, but also feeling disconnected from the social relationships they had built with their co-workers. In particular those who were required to shield during the first wave found this experience very psychologically isolating and lonely.

> *“Working from home, you don’t have any anyone to talk to that much…and there’s because we’re not face to face, you know, the human interaction is a bit lost… So there’s nobody really to talk to and nowhere to go but the house…the only thing I really miss is the interaction human interaction with my colleagues,”* (CRN)

#### Adaptability

##### Building confidence

Some nurses described reluctance from their colleagues or even themselves to use these new platforms. Many felt this reluctance was eventually overcome when they developed confidence, especially with support from IT (information technology), which was noted to be important. Initially, participants felt that they had always been told that working in this manner would be impractical, which made the experience quite daunting.

> *“Streamlining clinics made it evident they were doing things for years that didn’t need to be done; the resistance from doctors previously to do telephone clinics is now over because they have no choice but to do it,”* (Matron)

When it came time to rapidly implement virtual care policies, many found it surprising how easily they could work remotely when given the right resources and support. While there were initial adjustments to be made, the majority did feel confident in their use of virtual platforms. However, it was still felt that further training for professionals on the use of virtual care was needed.

> *“Being able to implement virtual clinics was possible because the IT team attitude went from ‘No we can’t do that’ to implementing everything you needed to make it happen,”* (Matron)

It was also noted that using aspects of remote access allowed them easier access to attend training, which could often be difficult to arrange if it involve nurses leaving the clinical areas – despite the clear benefit of staff participating in new and available training. Use of these technologies could therefore be used to help to solve certain logistical paradoxes often experienced by healthcare staff.

> *“I think that’s it’s the not having to go places to train and to have meetings. I think that’s, for me, a really good thing,”* (Sister)

##### Losing the human touch

The most voiced criticism of using virtual technologies in the nursing role was not being able to see patients in person. Some found it difficult to assess patients thoroughly through virtual care, and that if there was no video link then it could be quite impersonal and hard to build rapport with a patient. Lacking visual cues to how patients were feeling was difficult for some nurses who felt they often relied on these to gain a better understanding of their patient’s needs.

> *“You can’t really pick up nonverbal cues from people in the same way…that’s harder because you’re not firing on all cylinders with your ears and your eyes and watching body language and things. You’re just listening to a voice,”* (CNS)

Similarly, they felt it was still important to be able to see some patients face-to-face if there were safeguarding issues or mental health problems, as they needed to be able to check in person to more effectively gauge how they were. Participants were also concerned that patients may not be able to disclose their circumstances if they were home with family present.

> *“I know that when we think about safeguarding with our patients, there’s quite a few that we do want to clock eyes on and make sure that we see them or bring them in. But I guess that could be decided on a patient to patient basis,”* (CNS)

Nurses who were apprehensive using technology, and who often relied on face-to-face assessments and the personal touch when dealing with clients, there was a disconnect described in their interactions with their patients. Lacking the visual clues meant they had to rely more on what patients were telling them, though a positive of this may have been encouraging the patient to use their voice and empowered them to engage more in their care. This perceived distance and disconnect could be aggravated further due to technological issues, such as feeds lagging, freezing or the picture not being clear enough. This highlighted that while some found virtual care more accessible after building their confidence with the technologies, there were still some nurses who felt ill equipped and disconnected from their work.

> *“There’s a lot of things with checking out without them knowing it just by looking at them, how they move around the clinic room, how they carry themselves their mood, you can’t pick that up on a telephone clinic. So I’m probably missing some quite important things that I wouldn’t normally miss,”* (CNS)

## DISCUSSION

Our evaluation explored the experiences of nurses utilising virtual care and remote working during the pandemic in a single hospital, with the aim of identifying elements that could be adopted as common practice. We found that there were a number of positive and negative aspects associated with virtual care, which lends further credence to past findings that posit virtual care as a system which benefits some but not all. Nurses believed that virtual care impacted the experience of their patients. It was felt that provision of virtual care greatly improved the accessibility of healthcare for some, which also helped to lower the rate of missed appointments and allowed support systems to be more involved in joining in in the conversations with the medical team. However the inverse of this was also voiced, with virtual care potentially posing obstacles for accessing healthcare due to lacking adequate resources, confidence, skill or support. Nurses worried that moving toward virtual care would alienate some patients who enjoyed the social routine of attending hospital appointments and interacting with staff on a regular basis.

In terms of the impact on the nurse’s own roles, reactions were somewhat mixed. Some enjoyed the flexibility of remote working while others found working from home could lead to virtual fatigue. Some nurses felt that remote working increased their productivity and organisation, while others struggled to establish boundaries between work and home life, leading to feeling overburdened and stressed. The use of virtual care was seen to improve the level of interdisciplinary working, but inversely could lead to isolation from one’s own team as a consequence. Attitudes towards virtual care and remote working were somewhat ambivalent to begin with, and some felt resistance from their co-workers to fully engage; however, through exposure many found they gained confidence. A grievance that all nurses expressed to some degree was the loss of the human connection when not working face-to-face with patients. While some managed to positively adapt, others found they could not adapt their ways of normally assessing patients for virtual care. The somewhat dyadic findings of this evaluation are in line with previous research on virtual care, which state that it is a system which works very well for some people but not for everyone, which is vital to keep in mind when considering its future applications (Bashshur & Shannon, 2009, Hollander & Carr, 2020, Stokel-Walker, 2020).

As seen in the literature, virtual care is accredited with greatly improving access to care across various populations (Lilliecrap, Hunter & Goldswain, 2019; Murphy et al, 2020). Nurses believed that adopting virtual care during the pandemic was fundamental in maintaining services. It was felt that beyond the pandemic that use of virtual care could enable healthcare to be more equitable, and reach further communities, which has also been shown previously (Wosick et al, 2020). Surveys of virtual care during past and the current pandemics have shown high levels of acceptance by the general public (Azad et al, 2012, Sinha et al, 2020). Furthermore, given that the priority of most patients is the ability to be connected with relevant healthcare professionals, many have shown they are happy for this to take place virtually (RGCP, 2020).

This evaluation mirrored these findings, believing that virtual care significant improved access of care. It was also noted that DNA rates were lower and decreasing the amount of appointments which were missed was thought to positively contribute to shorter waiting lists. This is critical for patients with quickly deteriorating conditions, or those seeking a timely diagnosis (Murphy et al, 2020). Nurse found that reduced levels of missed appointments allowed for higher volumes of patients to be seen by the appropriate professional, thus benefiting the system as a whole (Lilliecrap, Hunter & Goldswain, 2019).

Despite this, there were fears of alienating subsections of the population and a sense that certain patients were at a higher risk of falling through the gaps. For some it was the use of virtual technology itself which was likely to be an obstacle for engaging. Patient’s reluctance or hesitancy to try new technologies has been previously linked to lower success for virtual care (Lilliecrap, Hunter & Goldswain, 2019). While for some it was just a case of practise to help build their virtual literacy skills, there were others who simply could not engage throughout the pandemic which was cause for concern. Previous research has shown that populations such as the elderly or those with autism are more likely to encounter obstacles when using virtual care which may act as a deterrent (Narasimha et al, 2017, Wosik et al, 2020). Identifying those most at risk of becoming lost in a system of care is necessary to ensure these patients continue to have access and engage with their treatment plans. Increasing the accessibility of virtual care could have a polarizing effect on patient’s social support and environment. Allowing patients to access clinics in the comfort of their own home has the advantage of creating safe and comfortable surroundings to maintain calm (Bashshur & Shannon, 2009) while also increasing the likelihood of having their family present. Historically, communication between healthcare staff and family is a source of contention (Newell & Jordan, 2015). Patients may not always take in all the information they are given in clinics and allowing family to be present to engage in real time with nurses has shown to improve the retention of information and benefit patient and family anxieties (Newell & Jordan, 2015). Families frequently experience periods of liminality and powerlessness in the wake of illness; this could be addressed by increasing collaboration between them and the healthcare team through virtual clinics (Clay & Parsh, 2016).

However, the inverse of this is also true; some patients may have a more complex home life or have surroundings in which they do not feel safe or comfortable to discuss their experiences. Some patients may be subject to safeguarding concerns and need to interact with nursing staff away from potentially harmful elements of their home life. Those who have children may also struggle to engage virtually if they have concerns over discussing illness around them (Bashshur & Shannon, 2009).

In terms of remote working, nurses expressed that the main benefit of this was the flexibility it provided. It was felt that having the option to work from home at least some of the week when the pandemic ended could be hugely beneficial in terms of productivity, stress levels and for personal circumstances such as finances and time spent commuting. Allowing nurses to work from home also overcomes the stress associated with hot desking and lack of available workspaces in clinical areas, as well as providing a calm and organised place in which to work. However, experiences of remote working, came with negative consequences arising from poor boundaries between work and home. Some expressed difficulty with creating a proper work life balance, which has been found previously in other studies of remote working (Chattopadhyay, Davies & Adhiyaman, Giurge & Bohns, 2020). Those who work remotely are still entitled to routine breaks and working only within their agreed hours. However, some felt managers and co-workers increased pressure to work harder or faster merely because they were working remotely. This has been shown previously and should be considered a target of work culture to be dismantled moving forward (Chattopadhyay, Davies & Adhiyaman, 2020).

A concern of remote working is that it may contribute to burnout, caused by this lack of balance, particularly in people’s failure to separate work and home life (Giurge & Bohns, 2020). Burnout can lead to feelings of emotional exhaustion, depersonalisation and a lack of accomplishment in one’s work, which in turn increases the risk of errors and poorer patient outcomes (Tawfik, Profit & Magenthaler, 2018). Pandemics greatly increase the likelihood of staff burnout in general, and wellbeing must be closely monitored to avoid a service wide burnout following its resolution (Hoffmann et al, 2020). However, the majority of participants reported enjoying the experience of working from home as it reduced time commuting and allowed them to spend more meaningful time with family. Recent research has found that when the ability to work remotely is an option, it acts as a protective factor mitigating provider burnout (Chattopadhyay, Davies & Adhiyaman, 2020, Hoffmann et al, 2020). It would therefore be reasonable to suggest that remote working is at its most effective when it is optional rather than mandatory, and that when staff have the freedom to choose onsite or remote working, or a mixture of both, they are most likely to be benefit from its protective potential. Given the threat of burnout to healthcare workers during a pandemic, finding avenues to strengthen their practises is critical (Hoffmann et al, 2020).

Virtual care seemed to bolster interdisciplinary teamwork while at the same time alienating individual team connections. Moving MDT clinics to a virtual format made them overall more accessible to a wider range of staff, and nurses benefited from having the perspectives of many different disciplines on patient care. Many felt this was a positive step towards the more holistic model that healthcare has been moving more towards (Stokel-Walker, 2020). However, nurses who were working remotely felt isolated from their team, particularly when it was mandatory due to shielding. Teamwork is a fundamental cornerstone of the nursing profession and a protective factor against poor mental health and experiences of burnout (Sharma & Clarke, 2014).

When nurses were removed from their teams completely, they were shown to experience more negative emotions than those who had the option of working remotely occasionally throughout the week. Some felt a perception from their colleagues that those working remotely had it easy, and experienced guilt at not being part of the frontline defence of the virus. This is similar to other studies done during the COVID-19 pandemic, which have shown that staff relegated to home working ran the risk of feeling isolated and under appreciated by their team, despite the work they still contributed (Chattopadhyay, Davies & Adhiyaman, 2020). When virtual team meetings were held more frequently it improved team morale, allowed for the maintenance of previous relationships, encouraged further bonding and allowed remote staff to still feel part of the team. It was also found that these meetings had the potential to incorporate some elements of socialising which could further improve mood for both onsite staff and remote staff alike.

Previous attempts to introduce new technologies into nursing roles has been met with resistance, due in part to a need to protect established routines which safeguard staff from the chaotic and ephemeral nature of their work (Sharma & Clarke, 2014). The introduction of new technology can often be viewed as threatening to established routines, which has been shown in nurse’s reactions to the introduction of virtual care in the past (Sharma & Clarke, 2014). However, virtual care has been used with some success by nurses in the United States over the last few years (Bashshur & Shannon, 2009). Prior to the pandemic there was considerable interest in developing digital health strategies in the UK, and a rapid shift was made over the course of the first and second wave (Murphy et al, 2020, Sinha et al, 2020). Even with advances there were still some who are more hesitant to accept the structural integration of virtual care (Bashshur & Shannon, 2009, Stokel-Walker, 2020). This was reflected in our evaluation, as some nurses stated that they could not adapt their skillset for the new medium. This was particularly true for those who felt they relied very much on in person contact to provide proper patient assessments, and who showed a lower level of adaptability to virtual care than some of their colleagues. The loss of the human connection has been shown as a concern for many medical staff when first introduced to virtual care (Sinha et al, 2020).

This should be held within the same thought continuum of the patients who are likely to fall through the gaps of virtual care – some nurses are confident and have the ability to adapt to change, but there will be those who will not have as great a reserve for change and will therefore struggle with wide scale disruptions (Sharma & Clarke, 2014). Nurses who are unconfident or have trouble adapting to new manners of working would need extra support to build their virtual literacy. Akin to past findings, virtual care is as much a ‘some but not all’ experience for staff as much as it is for patients.

### Limitations

The current evaluation has a number of limitations. First, this was secondary analysis of a wider evaluation; therefore virtual care and remote working were not the sole focus of the interviews. Interviews specific in this area may have included additional probing questions on the barriers and challenges specific to this. Despite this, these themes emerged organically during the interviews, and were explored by the interviewer due its apparent impact on nurse’s experiences during COVID-19. The wide scale and in-depth discussion of these themes warranted the authoring of this paper, rather than it being a subtheme in a larger evaluation. Second, this was a single centre evaluation and reflected the practices and decisions made in this one organisation. However, as a large inner city university hospital, the results may resonate with other organisations. Thirdly, only nursing staff were interviewed, so it does not reflect other professional groups who were using virtual care, e.g. medics and allied health professionals, or a large number of administrative and clerical staff who were required to work from home. It is important that their experience and perceptions are elicited to inform any future policy/guidance. Finally, while we present patient experience, this is through the perception of nurses. To fully capture the experience of those utilising healthcare during the pandemic it would be necessary to engage patients on how they found the experience of interacting with their treatment through virtual care. Despite these limitations, we were able to evaluate changes to service delivery in real time so we have an accurate recollection of nurse’s experiences of using virtual care and working from home. It also included nurses in a range of roles so presents multiple perspectives.

## CONCLUSION

The COVID-19 pandemic is still a dominant feature in the landscape of healthcare and is likely to be so for some time to come. In order to continue to provide optimal and consistent care to patients, while protecting them, their families and our healthcare workers, use of virtual care is an imperative step forward. However, moving towards business as usual it is clear that virtual remote access has added benefit for being integrated into the way we continue to engage our patients. It is likely there will be many epistemic changes post-pandemic, and a return to the way in which we once worked is highly unlikely. In order to embrace what is sure to become the new normal, becoming versed in the use of virtual care seems both progressive and highly pragmatic. Historical reservations around working remotely have been clearly disproven, with a wide varieties of jobs being shown to be possible offsite and approaching this with a level flexibility is key to not only maximising the way our staff are working during times of social distancing, but beyond this as well. The simultaneous increase in productivity and decrease in perceptions of stress, combined with the readily available forms of technology show that it is capable to not only move forward in how we deliver healthcare, but ultimately to expand in ways which only some years ago would have seemed impractical. Lessons learnt during the pandemic should not be merely restricted to emergency protocols but become long-term fixtures in how we think about the delivery of healthcare in the future.

### Implications for nursing practice

The use of digital technology is central to the NHS Long term plan in the UK (NHS, 2019) and COVID-19 has highlighted the needed for an integrative approach to nursing practice as we know it. Results from this evaluation emphasised a number of benefits of virtual care for nurses and patients, which should be considered when integrating virtual care into post-pandemic nursing practice. In addition, limitations or concerns around virtual care require attention and further investigation. Most notably, the need for a typology to facilitate decision making around appropriateness of virtual care verses face-to-face consultation for individual patients and situations. Training programs are also needed to support nurses in how best to delivery virtual care and stay connected with their patients.

After the first wave of the pandemic the NHS launched *We are the NHS: People plan for 2020/21* (NHS, 2020), which outlined what people working in the NHS could expect to “foster a culture of inclusion and belonging” (NHS, 2020, pg 3). The report outlined the strategy for caring for staff working in the NHS and one of the central recommendations for retaining staff was flexible working. Flexible working can be more easily accommodated in administration, Monday to Friday and non-clinical roles but can be more challenging for nurses who are working shifts and deliver patient care. When local policies are being developed for flexible working, this needs to be considered and flexible options offered, such as self-rostering. In addition, pilot training programs have been rolled out aimed at improving skills specific to delivering virtual care.

## Data Availability

No data are available

## Acknowledgements

We would like to thank the Nursing and Midwifery Leadership Team for commissioning and supporting the evaluation these data were derived from. We would also like to thank all the participants for giving us their time during the pandemic and for sharing so honestly about the impact of working in a pandemic.

